# Impact of Nocturnal Blood Pressure Patterns on BMI, Diabetes, and Cardiovascular Health: A Comprehensive Analysis

**DOI:** 10.1101/2025.10.02.25337214

**Authors:** Delowar Hossain, Nawal Kalid, Jashnoor Kaur Bhatti, Sidharth Singh Bhinder, Richard Siow, Yuichiro Yano, Raja B. Singh, Anmol Kapoor

## Abstract

Ambulatory Blood Pressure Monitoring (ABPM) provides valuable insights into a patient’s 24-hour blood pressure (BP) patterns, offering an enhanced assessment of cardiovascular health, particularly in relation to body mass index (BMI), diabetes, and menopausal status. This method provides a more comprehensive understanding of BP dynamics compared to traditional office-based measurements, assisting healthcare professionals with devising a more accurate assessment of a patient’s cardiovascular risk. ABPM is increasingly growing in adoption as a service in medical practices and pharmacies due to the numerous advantages. This retrospective cohort study analyzes 24-hour ABPM data from 5618 participants to investigate the relationship between nocturnal BP fluctuations found through ABPM with BMI, diabetes, hypertension in both male and female participants. It also investigates the relationship between nocturnal BP fluctuations and menopausal status in female participants. These conditions are studied to ultimately assess whether BP fluctuations are potentially connected to an increased risk of developing chronic conditions such as cardiovascular dysfunction. The ABPM nocturnal blood-pressure fluctuations are categorized within four distinct categories: normal dipping, non-dipping, reverse dipping, and extra dipping. The findings of this study show that 44.11% of participants exhibit hypertension, with most of these participants experiencing abnormal nocturnal blood pressure patterns. Increased BMI, particularly in overweight and moderately obese categories, was significantly associated with non-dipping patterns. In addition, the relationship between menopausal status for female participants and nocturnal dipping patterns, early morning hypertension and diabetes diagnoses are assessed. These findings underscore the importance of ABPM in detecting abnormal BP patterns and their associations with key health indicators, advocating for its broader integration in clinical settings.

## 1. INTRODUCTION

Hypertension affects millions of individuals worldwide and is a major contributor to morbidity and mortality [1–6]. It plays a major role in the development of heart conditions, such as acute myocardial infarction, stroke, transient ischemic attacks, kidney failure, retinal complications, premature death, and disability [7–9]. Hypertension is the most important cardiovascular risk factor and continues to be the most diagnosed in adult primary care. In Canada, according to 2017-2018 data from the Canadian Chronic Disease Surveillance System (CCDSS), about 25% of adults aged 20 years and older are living with diagnosed hypertension [10]. According to the National Center for Health Statistics (NCHS), nearly half of adults in the United States of America aged 18 and older have hypertension, with a prevalence of 47.7% (approximately 119.9 million individuals) between August 2021 to August 2023 [11].

Despite advancements in healthcare, traditional office-based blood pressure (BP) measurements are still predominately used. The issue with this method of BP measurement is that it often falls short in capturing the full range of BP fluctuations, particularly nocturnal hypertension which is elevated BP during sleep. This limitation can result in inaccurate diagnoses and missed opportunities for early intervention. The issue is further complicated by two phenomena:

- **White-coat hypertension:** Elevated BP readings in clinical settings but normal when measured outside of clinical settings [12].
- **Masked hypertension:** Normal BP readings during a clinic visit but elevated during daily activities [13].

These conditions expose the shortcomings of relying on single-point BP measurements in traditional healthcare settings, emphasizing the need for more comprehensive monitoring methods [14–15].

ABPM has established that out-of-office and home BP monitoring is superior to clinic BP monitoring [16–19]. A 24-hour blood pressure monitoring device is designed to be wearable and intended to continuously measure BP over a 24-hour period. Typical ABPM machines record BP readings at intervals of every 20-30 minutes [20]. Nocturnal hypertension has been closely associated with serious health problems such as cardiovascular disease (CVD), stroke, and chronic kidney disease [21–24]. Unlike daytime blood pressure, nocturnal hypertension often goes unnoticed in traditional clinical settings, therefore supporting ABPM as an essential tool for identifying individuals at risk and ensuring timely intervention.

Nocturnal blood pressure (BP) patterns offer valuable insights into an individual’s cardiovascular health and serve as important indicators of potential risks. These patterns are generally classified into four categories: normal dipping, non-dipping, reverse dipping, and extra dipping, which describe how nighttime BP levels compare to daytime levels. Non-dipping and reverse dipping patterns are particularly worrisome because they are closely linked to negative health outcomes such as cardiovascular disease, diabetes, and kidney dysfunction [25–27]. Non-dipping patterns indicate inadequate physiological recovery during sleep, placing extra strain on the cardiovascular system and increasing the risk of early-morning hypertension, which is a major trigger for cardiovascular events.

Body mass index (BMI) plays a significant role in influencing BP patterns [28–31]. Individuals who are overweight or obese are more likely to exhibit non-dipping or reverse dipping patterns, which further increase their cardiovascular risk [32–36]. These abnormal patterns are not only linked to the physical strain of excess weight on the cardiovascular system, but also to metabolic issues such as insulin resistance and systemic inflammation, both of which are common in obese individuals.

Diabetes diagnoses have been shown to significantly increase with the prevalence of hypertension diagnoses, serving as significant comorbidities [60–63]. Hypertension is more common in patients with diabetes, while diabetes is also more common in hypertensive patients than in the general population. Current scientific literature shows a possible causal relationship relating diabetes to affecting hypertension diagnoses [61]. BP patterns and their connections to diabetes diagnoses can serve as critical health indicators.

Menopausal status plays a significant role in shaping the risk of hypertension [37–39], with postmenopausal women being particularly susceptible to nocturnal hypertension and early-morning blood pressure (BP) surges [40–41]. The drop in estrogen levels during menopause disrupts key cardiovascular protective mechanisms, such as nitric oxide-mediated blood vessel relaxation, vascular flexibility, and inflammation regulation. This hormonal shift leads to increased vascular stiffness, higher peripheral resistance, and reduced endothelial function, all of which contribute to abnormal BP regulation.

As a result, postmenopausal women are more likely to exhibit non-dipping or reverse dipping nocturnal BP patterns [42–43]. These patterns indicate that blood pressure does not adequately decrease during sleep, signaling insufficient recovery for the cardiovascular system. This lack of recovery is strongly linked to an increased risk of serious health conditions such as stroke, coronary artery disease, and chronic kidney disease. Early-morning BP surges are more pronounced in postmenopausal women and occur during the critical transition from rest to activity, which is driven by heightened sympathetic nervous system activity and reduced baroreceptor sensitivity. These surges significantly raise the likelihood of acute cardiovascular events, including heart attacks and strokes.

ABPM is a powerful tool for tracking 24-hour BP fluctuations, including nighttime patterns, making it essential for effective hypertension management [44–46]. By identifying abnormal nocturnal BP profiles, ABPM enables healthcare providers to implement targeted interventions, such as personalized medication plans and lifestyle modifications. This comprehensive approach not only improves the accuracy of hypertension diagnosis but also enhances risk assessment and treatment strategies, ultimately helping to reduce the burden of cardiovascular diseases. Additionally, ABPM distinguishes between white-coat hypertension and masked hypertension, preventing misdiagnoses and ensuring patients receive appropriate care [47–48].

### Study Objectives

1. This study examines the association between nocturnal BP fluctuations, BMI, and diabetes to link these parameters to chronic conditions like cardiovascular and renal dysfunctions.
2. Examining data from 5618 participants, the study categorized nocturnal BP patterns into four categories: normal dipping, non-dipping, reverse dipping and extra dipping.
3. The study further examines any potential association between menopausal status and nocturnal BP patterns in female participants, aiming to assess the implications on their cardiovascular health.

This study highlights the unique ability of ABPM to uncover nocturnal BP patterns and their connections to critical health indicators such as BMI, diabetes, and menopausal status. By providing comprehensive data on 24-hour BP fluctuations, ABPM offers a transformative approach to managing hypertension. The findings emphasize the importance of integrating ABPM into routine clinical practice to enable earlier detection, more accurate diagnoses, and personalized treatment strategies. This approach has the potential to improve patient outcomes and reduce the prevalence of complications associated with high blood pressure, contributing to better cardiovascular health on a broader scale.

## 2. METHODS

### 2.1 Blood Pressure Measurement Device

The BPAro is a FDA and Health Canada approved 24-hour ABPM device (shown in Figure 1) that measures blood pressure at the brachial artery level [49]. ABPM devices are designed for portability, enabling the collection of blood pressure readings over a full 24-hour period at regular intervals. These devices are essential for assessing nocturnal blood pressure dipping patterns, which are critical for predicting and evaluating a patient’s health. A full description of the device is available on the BPAro’s website [50].

**Figure 1.**
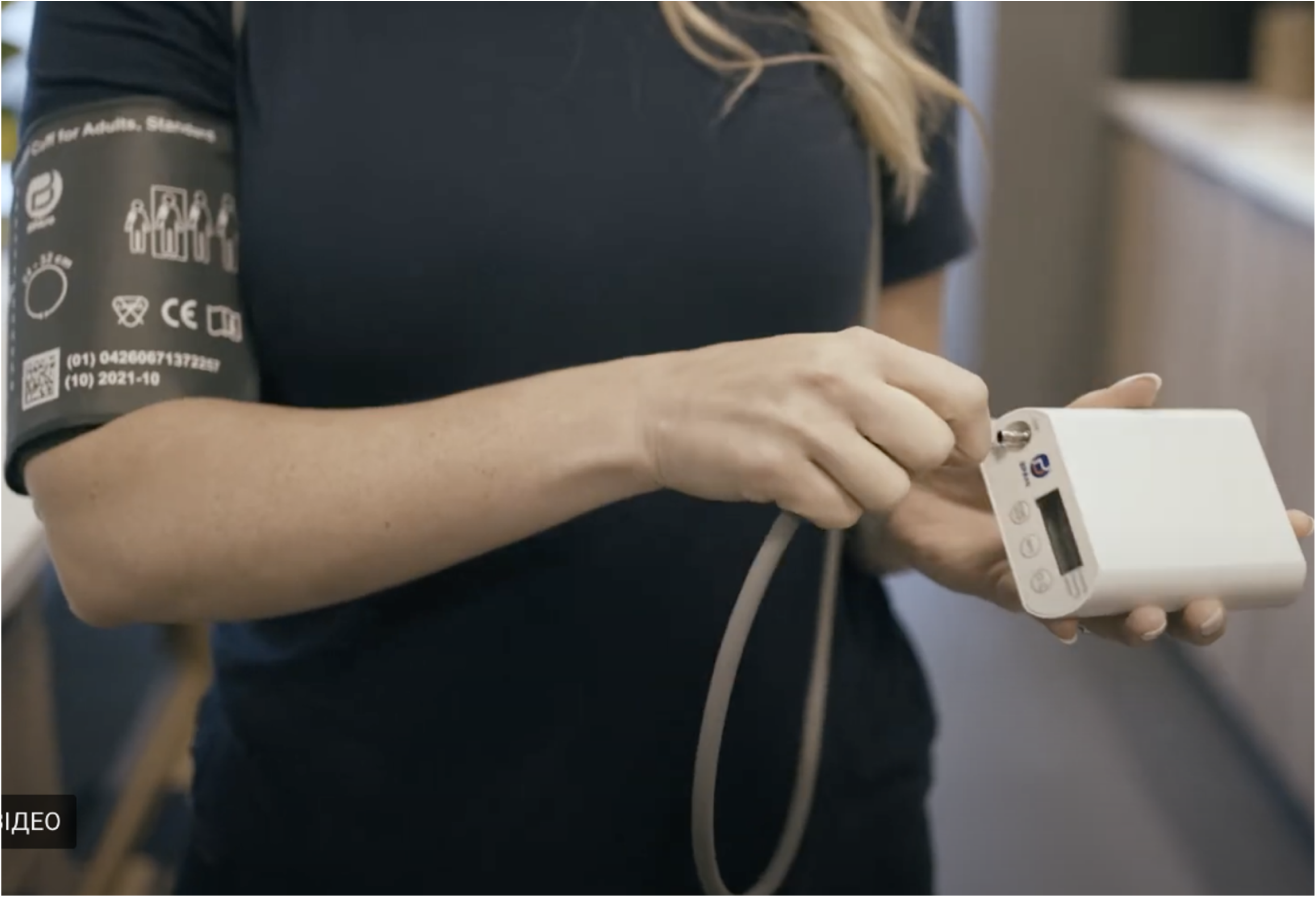
24-hour Ambulatory Blood Pressure Monitor (ABPM), also as BPAro.

Each patient was professionally fitted with appropriately sized cuffs to ensure measurement accuracy. The ABPM device was provided in a wearable pouch with an attached shoulder strap to stabilize the monitor. The hose connecting the cuff to the monitor was secured around the shoulders to minimize movement and maintain accuracy. The BPAro records BP readings approximately every 20–30 minutes during the monitoring period.

Reports generated by the BPAro were automatically processed using its supplementary software. These reports were sent to the patient’s physician for review and verification by a licensed cardiologist. Additionally, patients were able to access their reports directly through the BPAro app once the 24-hour ABPM test was complete and the data had been processed.

### 2.2 Study Design and Participants

This retrospective study analyzed a total of 5618 patient reports generated by BPAro, in collaboration with Advanced Cardiology Consultants and Diagnostics Inc. [80]. The data was collected from both treated and untreated patients between the ages of 18 to 74 years who underwent ABPM testing for various clinical indications between October 2022 and July 2023. The mean age of the study population was 52.72 years. Among the participants, 55.38% were male, and 44.62% were female.

Patients included in the study were referred by physicians for 24-hour ABPM testing due to pre-existing clinical indications. The BPAro ABPM device recorded blood pressure measurements approximately every 30 minutes over a 24-hour period. The reports generated by the BPAro supplementary software were automatically emailed to the referring physicians and subsequently verified by certified cardiologists.

This study was reviewed and approved by the Health Research Ethics Board of Alberta (HREBA) – Community Health Committee (CHC). All participants gave their written informed consent before performing the assessment.

### 2.3 Data Collection and Parameter Definitions

This study collected various patient parameters, including body mass index (BMI), 24-hour ABPM readings (daytime and nocturnal blood pressure), hypertension diagnosis status, diabetes diagnosis status, heart rate, age, sex, and early morning hypertension status.

Hypertension status was determined based on the Canadian Journal of Cardiology 2017 guidelines [20]. A mean awake daytime ABPM reading of ≥135/85 mmHg was classified as hypertension, while readings below this threshold (<135/85 mm Hg) indicated non-hypertensive status. Nocturnal blood pressure readings, along with specific before- and after-waking measurements, were programmed to align with each patient’s regular sleep and wake schedule, as entered the BPAro App software to assess early morning hypertension.

Patient BMI values as well as self-reported diabetes diagnoses were entered into the BPAro App software before the 24-hour ABPM study began. Premenopausal and postmenopausal status for female participants were also determined using the collected sex and age data. Women under the age of 50 were classified as premenopausal, while those aged 50 and above were categorized as postmenopausal [51].

### 2.4 Theoretical Framework: Categories of Nocturnal Blood Pressure Dipping

A theoretical framework was used to classify nocturnal blood pressure patterns into four categories: normal dipping, non-dipping, extra dipping, and reverse dipping (Figure 2).

- **Normal Dipping**: A decrease in mean nocturnal systolic blood pressure by 10% to 20% compared to the mean daytime systolic blood pressure.
- **Non-Dipping**: A decrease of less than 10% in mean nocturnal systolic blood pressure, without any increase, relative to the mean daytime systolic blood pressure.
- **Extra Dipping**: A decrease of more than 20% in mean nocturnal systolic blood pressure compared to the mean daytime systolic blood pressure.
- **Reverse Dipping**: An increase in mean nocturnal systolic blood pressure instead of a decrease, when compared to the mean daytime systolic blood pressure.

**Figure 2.**
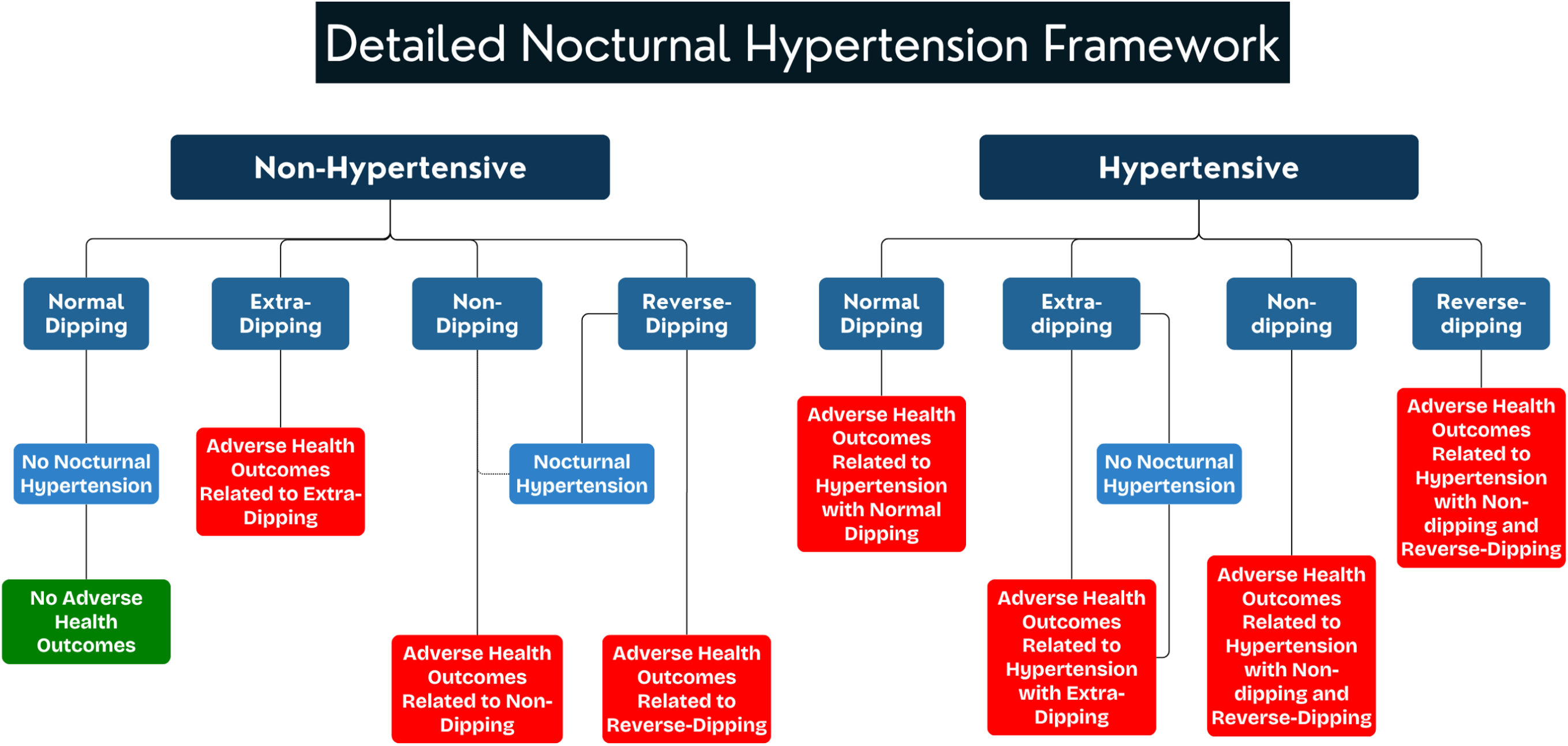
Diagnostic Framework for Nocturnal Hypertension: Categorization of Nocturnal Blood Pressure Dipping Patterns.

This framework begins by dividing individuals into two overarching groups: hypertensive and normotensive.

- **Hypertensive**: Hypertension is defined as a mean daytime blood pressure of 135/85 mmHg or higher, as measured by an automated ABPM device.
- **Non-Hypertensive**: Non-hypertension is defined as a mean daytime blood pressure below 135/85 mmHg.

These categories provide the foundation for further stratification into specific risk groups. While initial classification as hypertensive or non-hypertensive often occurs during an in-office visit, ABPM testing refines this classification by identifying nocturnal blood pressure dipping patterns.

**Nocturnal hypertension** is defined as a mean nocturnal blood pressure of 120/70 mm Hg or higher [52], which can occur in both hypertensive and non-hypertensive individuals. Since daytime blood pressure readings are currently the primary criterion for diagnosing hypertension, using ABPM is more beneficial as it provides more precise blood pressure data, improving diagnostic accuracy.

**Early Morning Hypertension** is defined as BP readings exceeding 135/85 mmHg during the first two hours after waking [64].

Non-hypertensive patients with a normal dipping pattern are generally at low risk for adverse cardiovascular outcomes whereas hypertensive patients with a normal dipping pattern are still at risk for adverse events (Figure 2). Extra dipping patterns in both normotensive and hypertensive individuals may signal risks such as blood pressure instability, fainting, or orthostatic hypotension [53]. Non-dipping and reverse dipping patterns, regardless of hypertension status, are linked to adverse outcomes like cardiovascular disease, autonomic dysfunction, kidney issues, diabetes, glucose intolerance, and obstructive sleep apnea [54]. Reverse dipping is particularly associated with a heightened risk of severe cardiovascular events and mortality [53].

By classifying patients into these nocturnal blood pressure patterns, ABPM enhances the ability to identify at-risk individuals and tailor treatment strategies more effectively.

### 2.5 Body Mass Index (BMI)

Body mass index (BMI) is a commonly used tool for classifying individuals based on their weight relative to their height. It is divided into six categories: severely underweight, underweight, normal weight, overweight, moderately obese, and severely obese. According to the World Health Organization (WHO) [55, 57], the classifications are as follows:

- Severely Underweight: BMI less than 16.5 kg/m²
- Underweight: BMI between 16.5 and 18.4 kg/m²
- Normal weight: BMI between 18.5 and 24.9 kg/m²
- Overweight: BMI between 25 and 29.9 kg/m²
- Moderately obese: BMI between 30 and 34.9 kg/m²
- Severely obese: BMI of 35 kg/m² or higher

These BMI categories play a key role in assessing the risk of various health issues. Overweight and obese individuals are at a higher risk of developing conditions such as cardiovascular disease, diabetes, and hypertension. By providing a standardized and practical method for evaluating weight status, BMI helps in population-level health assessments and risk stratification, guiding interventions to address these health concerns.

### 2.6 Statistical Analysis

The statistical analysis of the collected data was conducted in two main stages: data compilation and statistical evaluation. Data obtained using the BPAro ABPM App was systematically compiled and organized into a master table using Microsoft Excel. Data analysis was performed using Python version 3.12 [56] for Windows.

Descriptive statistics were used to summarize the data, including calculations of the mean and standard deviation for quantitative variables, as well as frequency and proportion for categorical variables. Independent t-tests were conducted to compare the means and other parameters between two groups.

The mean, which represents the average value and indicates the central tendency of the data, was calculated by summing all data points and dividing them by the total number of points. The standard deviation, a measure of how much the data varies around the mean, was calculated as the square root of the variance. These statistical tools were instrumental in analyzing and comparing nocturnal blood pressure dipping patterns and their associated risk factors, providing valuable insights into the data.

The formula used to calculate mean (𝑥) is:

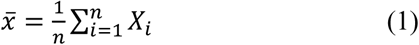

Where 𝑋_𝑖_ is the *i-*th sample, *n* is the total number of samples.

The formula for calculating the standard deviation (σ) is:

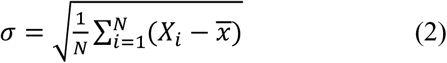

where σ is the standard deviation, N is the total number of samples, 𝑋_𝑖_ is the *i*-th sample value, and 𝑥 is the mean of the dataset.

Pearson’s Chi-Square Test: The Pearson’s Chi-Square Test, denoted as 𝜒^2^, is a statistical method used to determine whether there is a significant difference between an observed set of results and the expected outcomes. It evaluates how well the observed data matches the expected distribution under a given hypothesis.

The formula for calculating the Chi-Square statistics is:

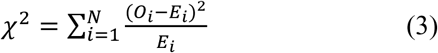

Where 𝜒^2^is the Chi-Square statistic, 𝑂_𝑖_ represents *i*-th sample of the observed frequency, 𝐸_𝑖_ represents the *i*-th sample of the expected frequency, *N* presents the total number of samples, and ∑ indicates the summation over all samples. This test is commonly used in hypothesis testing to access relationships between categorical variables in contingency tables.

## 3. EXPERIMENTAL RESULTS

A total of 5618 study subjects were included in this analysis. Among the participants, 44.62% were female, and 55.38% were male (Table 1). The mean age of the participants was 52.72 years, with a standard deviation of ±13.17 years.

**Table 1.**
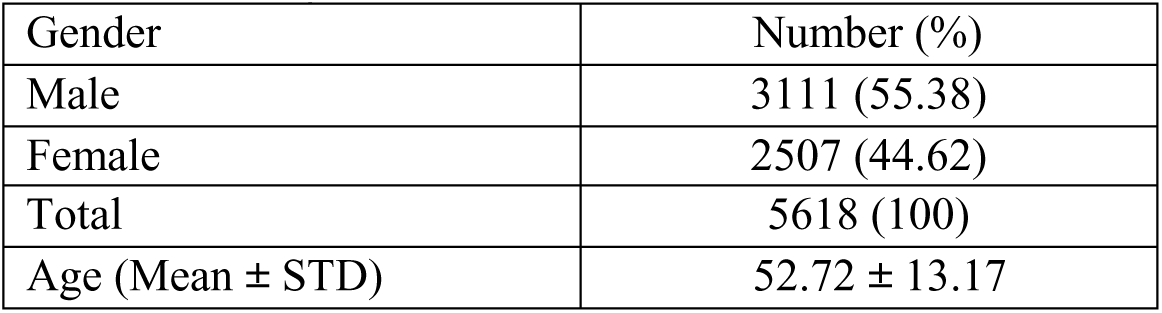
Demographic Information of 7925 patients

A sample patient’s 24-hour ABPM Data is shown in Figure 3. The shaded area represents the sleep period, during which 22 blood pressure readings were recorded. A total of 31 measurements were taken during the awake period. The overall average blood pressure was 119/79 mmHg, with an awake average of 122/81 mmHg and a sleep average of 114/75 mmHg. This reflects a 6.6% reduction in nocturnal systolic and 7.4% of nocturnal diastolic blood pressure. The average pulse rate was 81 bpm.

**Figure 3.**
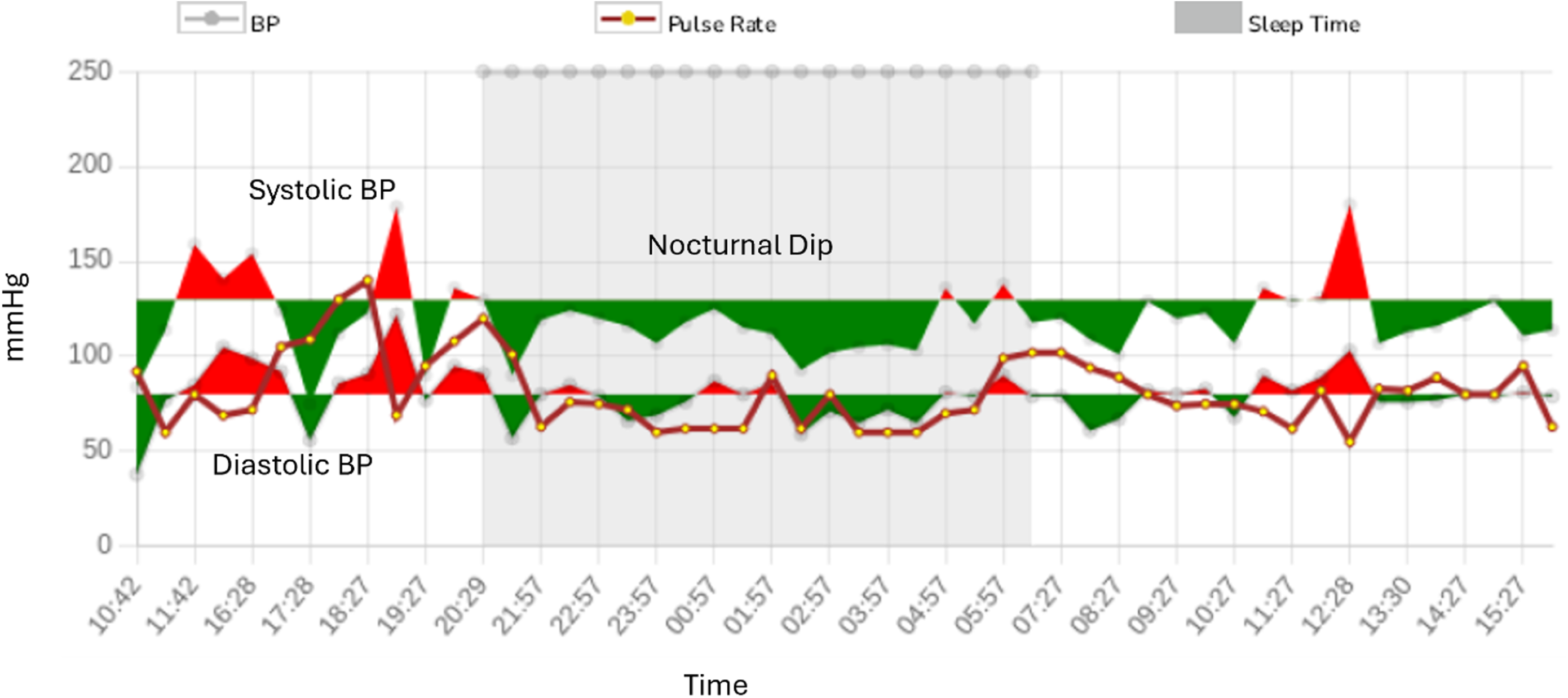
Sample of a 24-hour ABPM Data of a patient. BP = blood pressure; bpm = beats per minute.

To normalize the data within each group, the proportion of the total was calculated by dividing each value by the sum of all values in the group and multiplying by 100, yielding the percentage of the total. This is a retrospective cohort study, therefore there is no guarantee of equal group sizes. This approach facilitates effective comparison between groups, particularly when group sizes or total number of data points in a specific category differ across parameters.

### 3.1 Hypertension Diagnoses in Relation to Nocturnal Dipping Patterns

The correlation between hypertension status and nocturnal dipping patterns is found in Figure 4. Among the normal dipping category, 44.11% of participants are hypertensive while 55.89% are non-hypertensive. The p value is equal to 0.0017 which indicates that these findings are statistically significant (p < 0.05). The standard error is equal to 1.19 in this category.

**Figure 4.**
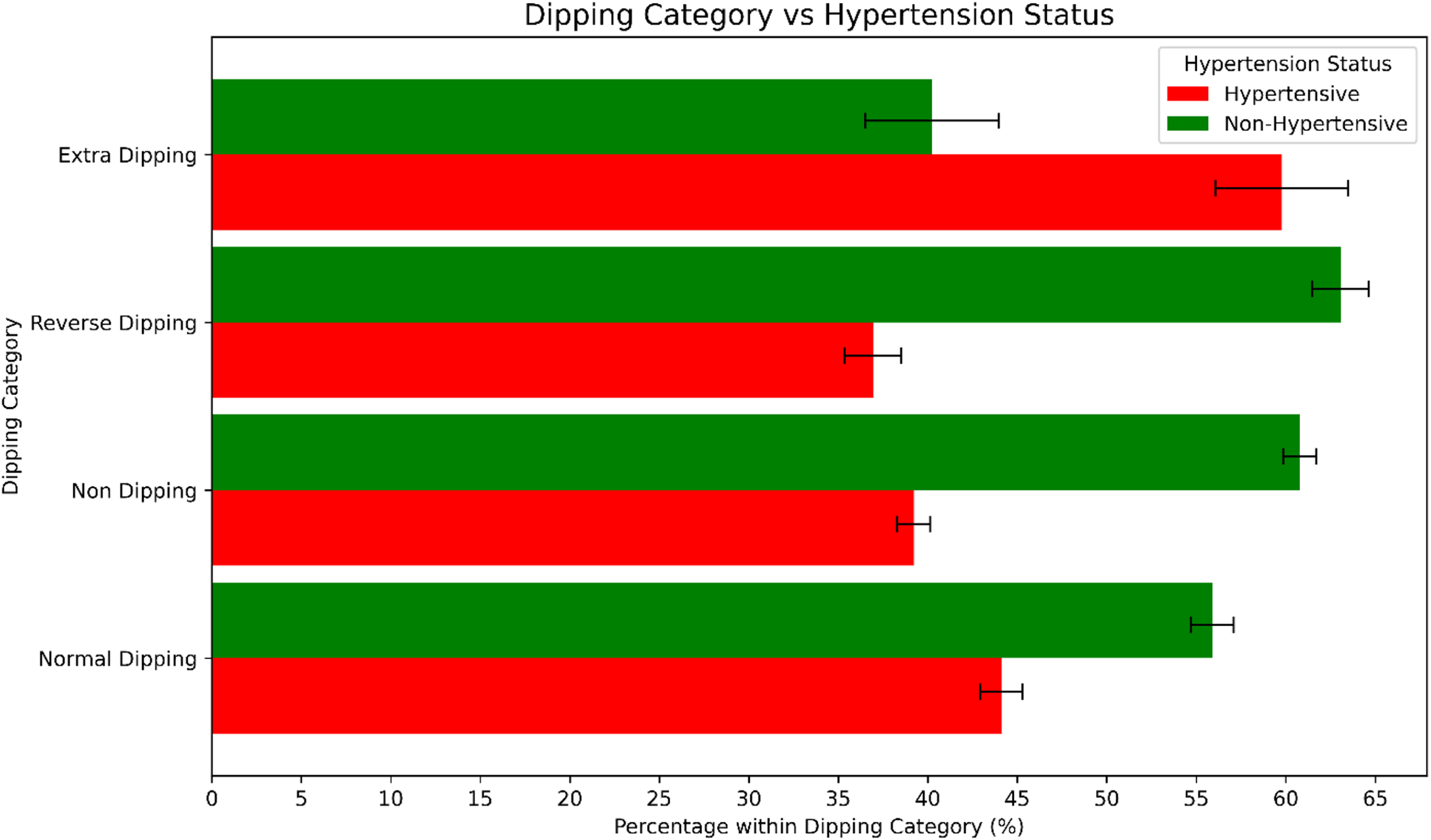
Hypertensive and non-hypertensive individuals evaluated amongst nocturnal dipping categories.

Within the non-dipping category, 39.23% of participants are hypertensive, and 60.77% are non-hypertensive. The difference is statistically significant (p < 0.05) with a p value equal to 0.0081. The standard error for both groups in this category is 0.92.

The reverse dipping category has 36.94% hypertensive participants and 63.06% non-hypertensive participants with a p value of 0.0070. This difference is statistically significant (p < 0.05) and the standard error for both groups in this category is 1.59.

Among participants in the extra dipping group, 59.77% are hypertensive while 40.23% are non-hypertensive. This difference is not statistically significant (p > 0.05) with a p value equal to 4.71. The standard error for both groups is 3.71.

### 3.2 Hypertension in Relation to BMI

The correlation between hypertension status and BMI is found in Figure 5(a). Among the severely underweight category (BMI < 16.5), 23.81% of participants are hypertensive whereas 76.19% of participants are non-hypertensive, with a p value of 0.17. The p-value indicates that the results are not statistically significant (p > 0.05). The standard error for groups is 9.29.

**Figure 5(a).**
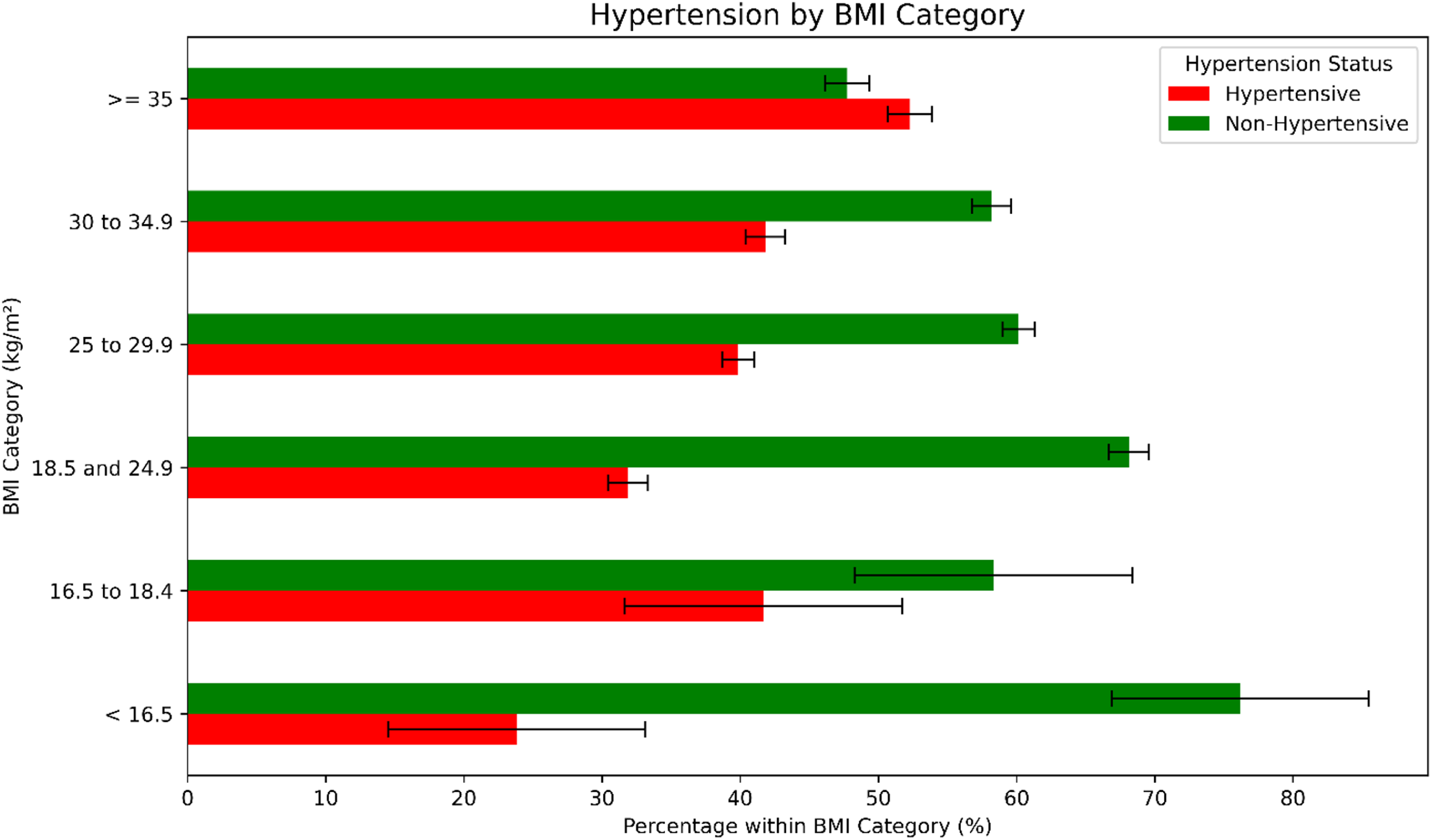
Hypertensive and non-hypertensive individuals evaluated amongst BMI categories.

Within the underweight category (16.5 to 18.4), 41.67% of participants are hypertensive and 58.33% are non-hypertensive, with a p value of 1.0. The p-value indicates that the results are not statistically significant (p > 0.05) and the standard error for both groups is 10.06.

For the participants in the normal weight category (18.5 to 24.9), 31.87% hypertensive and 68.13% of are non-hypertensive, with a p value of 3.51e-11. This difference is strongly statistically significant (p < 0.05). The standard error in this group is 1.44.

In the overweight category (25 to 29.9), 39.85% of participants are hypertensive and 60.15% are non-hypertensive, with a p value of 0.25 and a standard error of 1.17. The p-value indicates that the results are not statistically significant (p > 0.05).

Among the moderately obese category (30 to 34.4), 41.80% of participants are hypertensive, and 58.20% are non-hypertensive. The p value is equal to 0.54, therefore the results are not statistically significant (p > 0.05). The standard error for both groups is 1.42.

A statistically significant difference (p = 5.55e-15, p < 0.05) is observed in the severely obese category (BMI > 35), where 52.27% of participants are hypertensive and 47.73% are non-hypertensive. The standard error for groups is 1.60.

### 3.3 Nocturnal BP Patterns in Relation to BMI

The correlation between hypertension status and BMI is found in Figure 5(b). Among the severely underweight category, 28.57% of participants have normal dipping nocturnal BP patterns (Standard Error (SE) = 9.86), 47.62% experience non dipping (SE = 10.90), 19.05% experience reverse dipping (SE = 8.57) and 4.76% experience extra dipping (SE = 4.65). The p value is equal to 0.95, therefore the difference is not statistically significant (p > 0.05).

**Figure 5 (b).**
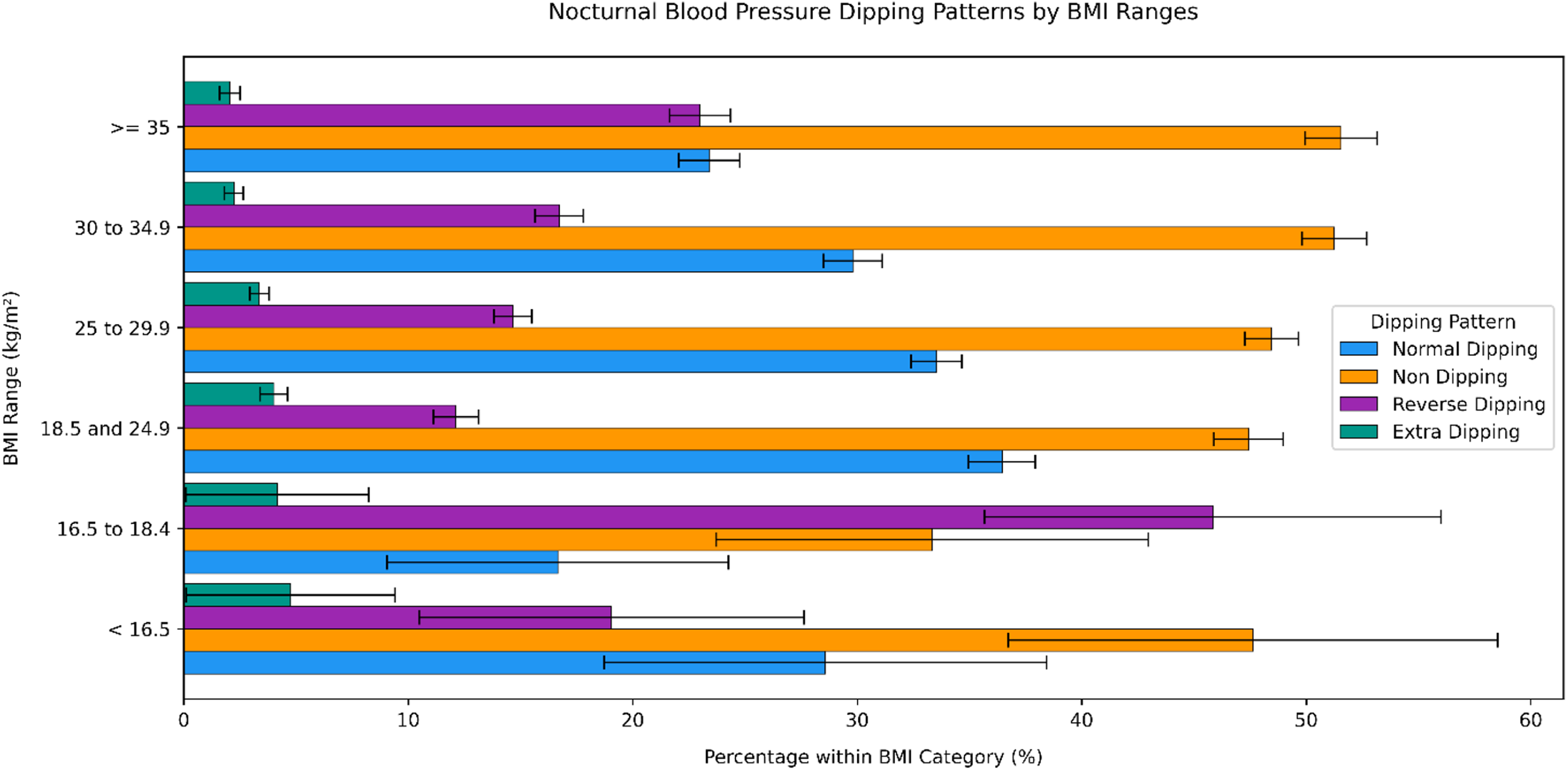
Normal dipping, non-dipping, reverse dipping and extra dipping participant nocturnal BP patterns evaluated amongst BMI categories.

Within the underweight category, 16.67% of participants are normal dippers (SE = 7.61), 33.33% are non-dippers (SE = 9.62), 45.83% are reverse dippers (SE = 10.17) and 4.17% are extra dippers (SE = 4.08). The p value is equal to 0.001, therefore the difference is statistically significant (p < 0.05).

For the participants in the normal weight category, 36.45% experience normal dipping (SE = 1.49), 47.42% experience non dipping (SE = 1.54), 12.12% experience reverse dipping (SE = 1.01) and 4.01% experience extra dipping (SE = 0.61). The p value is equal to 4.55e-07, therefore the difference is strongly statistically significant.

For the participants in the overweight category, 33.52% experience normal dipping (SE = 1.13), 48.46% experience non dipping (SE = 1.19), 14.65% experience reverse dipping (SE = 0.84) and 3.36% experience extra dipping (SE = 0.43). The p value is equal to 0.009, therefore the difference is statistically significant.

For the participants in the moderately obese category, 29.80% experience normal dipping (SE = 1.32), 51.24% experience non dipping (SE = 1.44), 16.72% experience reverse dipping (SE = 1.07) and 2.24% experience extra dipping (SE = 0.43). The p value is equal to 0.16, therefore the difference is not statistically significant.

For the participants in the severely obese category, 23.40% experience normal dipping (SE = 1.36), 51.55% experience non dipping (SE = 1.60), 22.99% experience reverse dipping (SE = 1.35) and 2.06% experience extra dipping (SE = 0.46). The p value is equal to 6.77e-13, therefore the difference is strongly statistically significant.

### 3.4 Diabetes and the Association with Hypertension

The correlation between diabetes and hypertension is found in Figure 6(a). Among the hypertensive category, 82.41% of participants are non-diabetic and 17.59% are diabetic, with a p value of 0.001 and a standard error of 0.79. The difference is statistically significant (p < 0.05).

**Figure 6(a).**
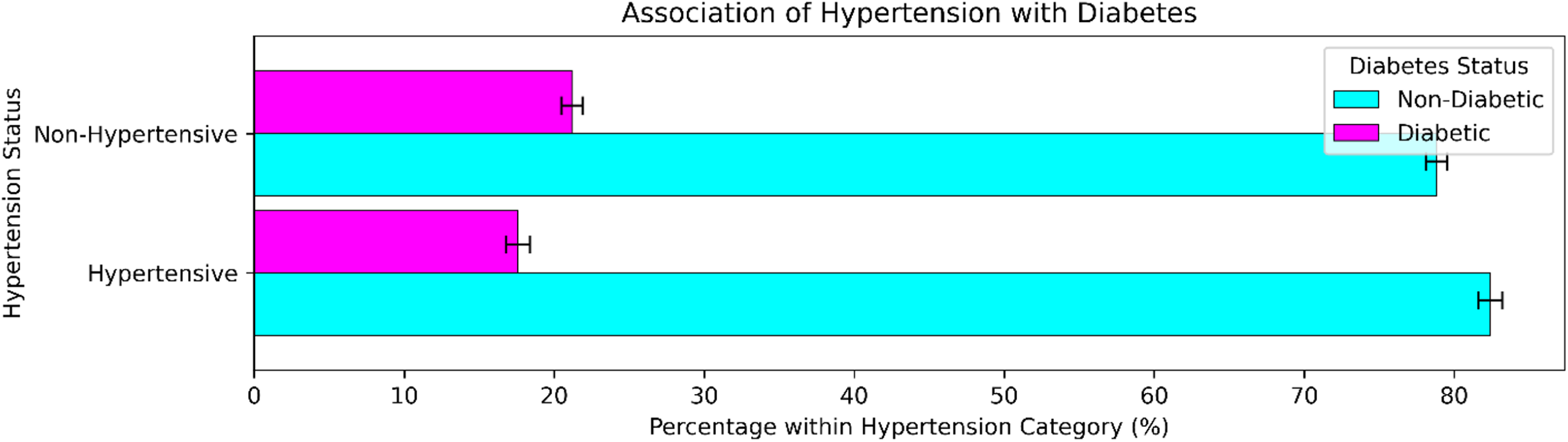
Diabetic and non-diabetic individuals evaluated amongst hypertensive status.

In the non-hypertensive category, 78.82% of participants are non-diabetic and 21.18% are diabetic with a p-value of 0.001 and a standard error of 0.71. The difference is statistically significant (p < 0.05).

### 3.5 Diabetes and the Association with BMI

The correlation between diabetes and BMI is found in Figure 6(b). There is no available diabetic data within the severely underweight category (BMI < 16.5).

**Figure 6(b).**
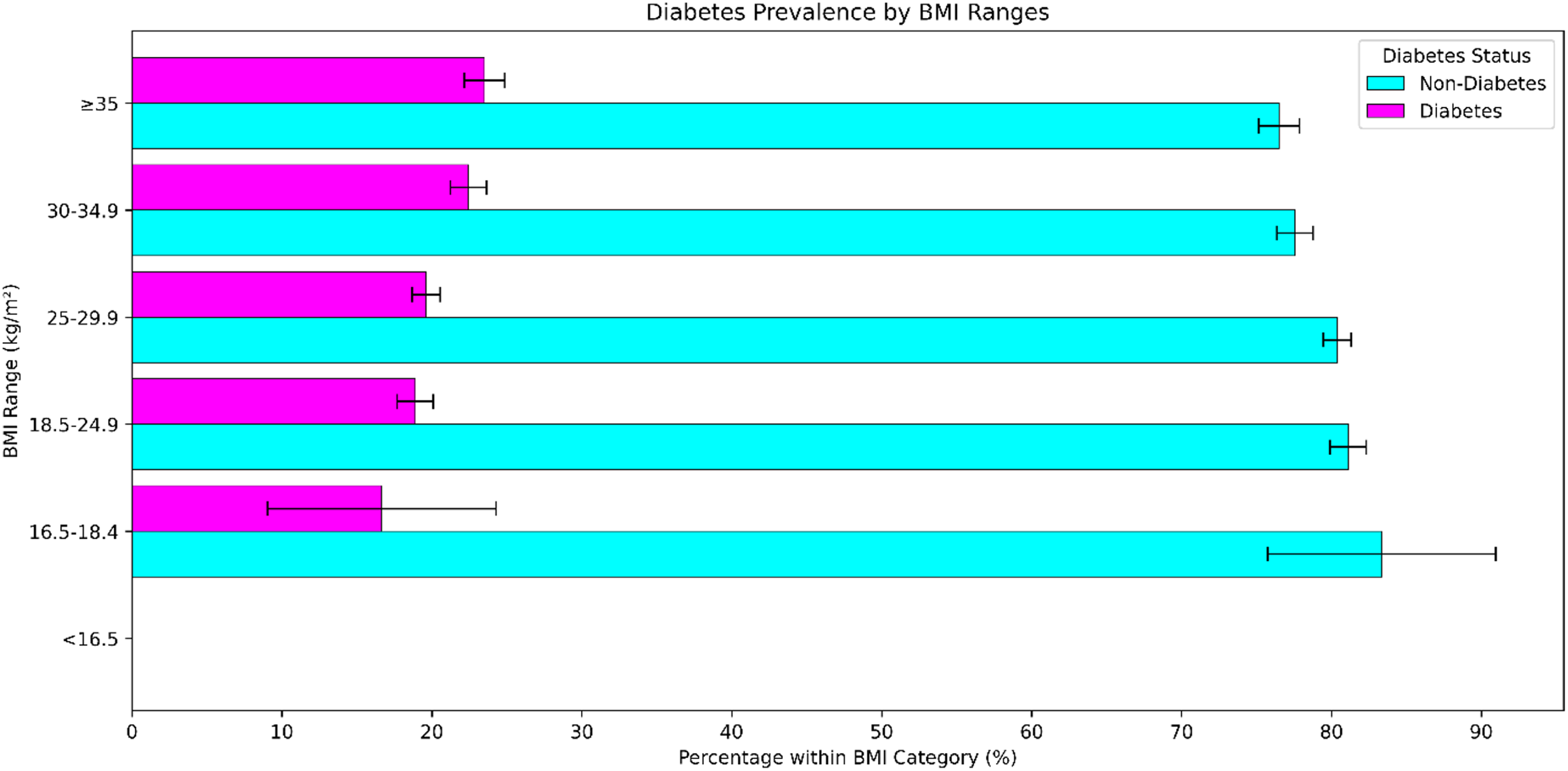
Diabetic and non-diabetic individuals across BMI categories.

Within the underweight category (16.5 to 18.4), 83.33% of participants are non-diabetic and 16.67% are diabetic. The p value is equal to 0.80 therefore these findings are not statistically significant (p > 0.05). The standard error is 7.61.

Among the normal weight category (18.5 to 24.9), 81.11% of participants are non-diabetic and 18.89% are diabetic with a p value equal to 0.082. Therefore, the difference is not statistically significant (p > 0.05). The standard error is 1.21.

In the overweight category (25 to 29.9), 80.39% of participants are non-diabetic and 19.61% are diabetic with a p value of 0.11 which indicates the difference is not statistically significant (p > 0.05). The standard error is 0.95.

Among the moderately obese category (30 to 34.9), 77.57% of participants are non-diabetic and 22.43% are diabetic. The p value is equal to 0.14, therefore the difference is not statistically significant (p > 0.05). The standard error is 1.20.

A statistically significant difference (p = 0.03, p < 0.05) is observed in the severely obese category (BMI > 35), where 76.49% of participants are non-diabetic and 23.51% are diabetic. The standard error for groups is 1.36.

### 3.6 Diabetes and the Association to Nocturnal BP Patterns

The correlation between diabetes and nocturnal dipping patterns is found in Figure 6(c). In the normal dipping category, 82.22% of participants are non-diabetic and 17.78% are diabetic. The p value is equal to 0.02, indicating that these findings are statistically significant (p < 0.05). The standard error is 0.92.

**Figure 6(c).**
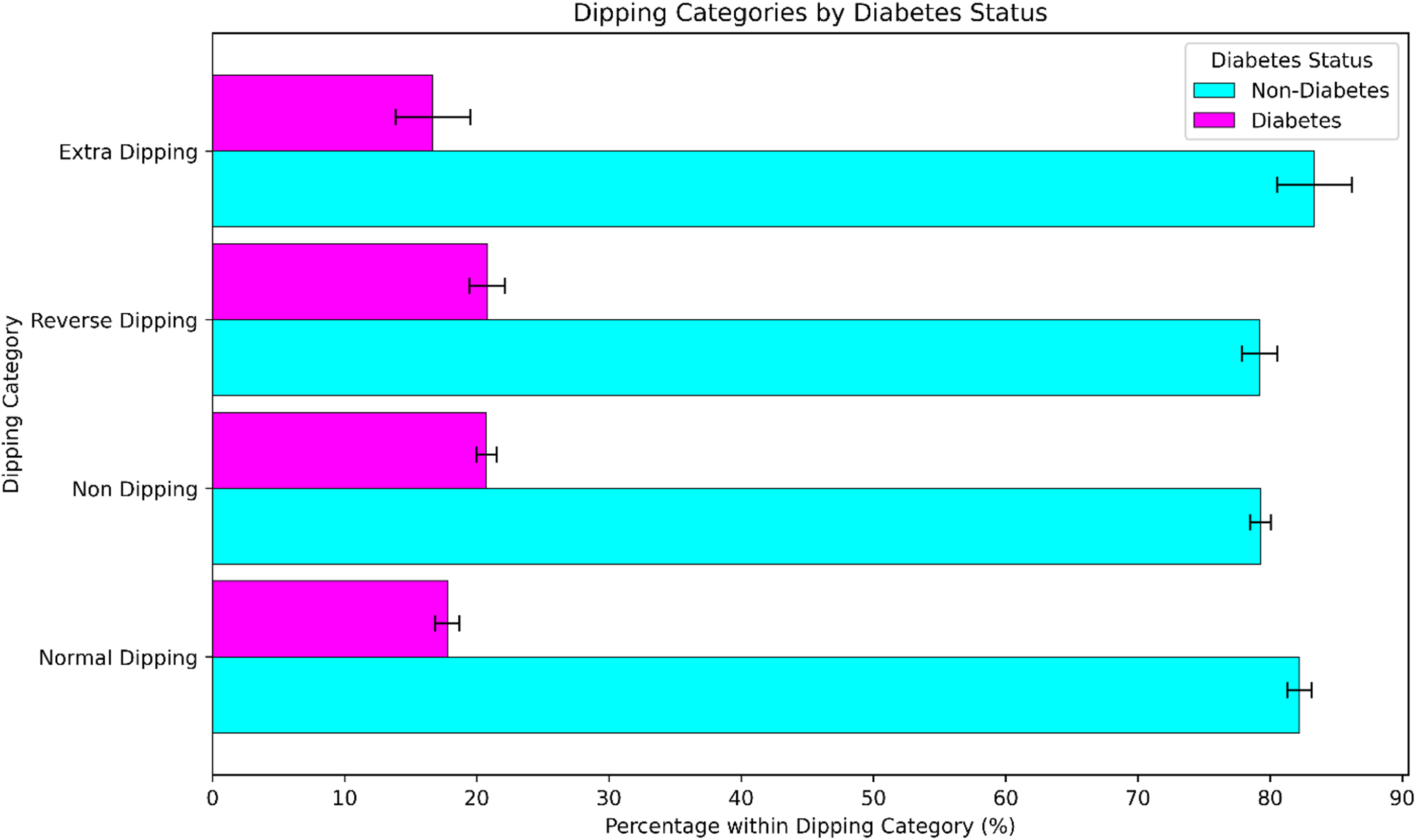
Diabetic and non-diabetic individuals evaluated amongst nocturnal BP dipping patterns.

For the participants in the non-dipping category, 79.28% are non-diabetic and 20.72% are diabetic. The p value is equal to 0.06 therefore the difference is not statistically significant (p > 0.05). The standard error is 0.77.

In the reverse dipping category, 79.20% of participants are non-diabetic and 20.80% are diabetic with a p value of 0.38. The difference is not statistically significant (p > 0.05). The standard error is 1.34.

For the participants in the extra dipping category, 83.33% are non-diabetic and 16.67% are diabetic with a p value of 0.35. The difference is not statistically significant (p > 0.05) and the standard error is 2.83.

### 3.7 Menopause and the Association with Hypertension in Female Participants

The correlation between menopausal status and hypertension is found in Figure 7(a). In the hypertensive category, 61.03% of women are post-menopausal and 38.97% of women are pre-menopausal. The p value is equal to 0.24, therefore the difference is not statistically significant (p > 0.05). The standard error is 1.55.

**Figure 7(a).**
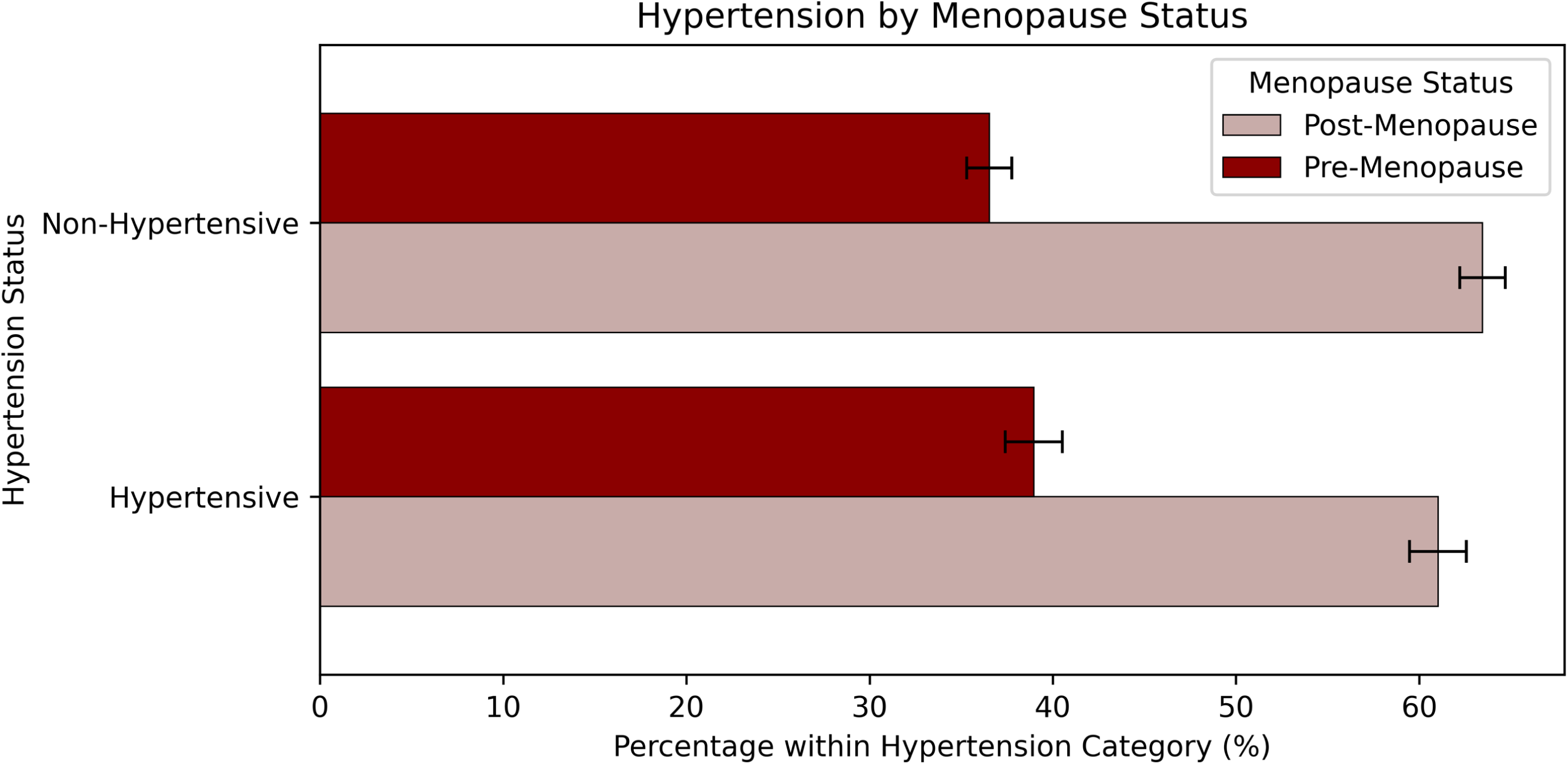
Pre-menopausal and post-menopausal women evaluated amongst hypertensive status.

In the non-hypertensive category, 63.46% of female participants are post-menopausal women and 36.54% of female participants are pre-menopausal women. The p value is 0.24, therefore the difference is not statistically significant (p > 0.05). The standard error is 1.24.

### 3.8 Menopause and the Association with Diabetes

The correlation between menopausal status and diabetes is found in Figure 7(b). Among the pre-menopausal category, 84.04% of female participants are non-diabetic and 15.96% of female participants are diabetic. The p value is equal to 0.0001 which indicates that the difference is statistically significant (p < 0.05). The standard error is 1.19.

**Figure 7(b).**
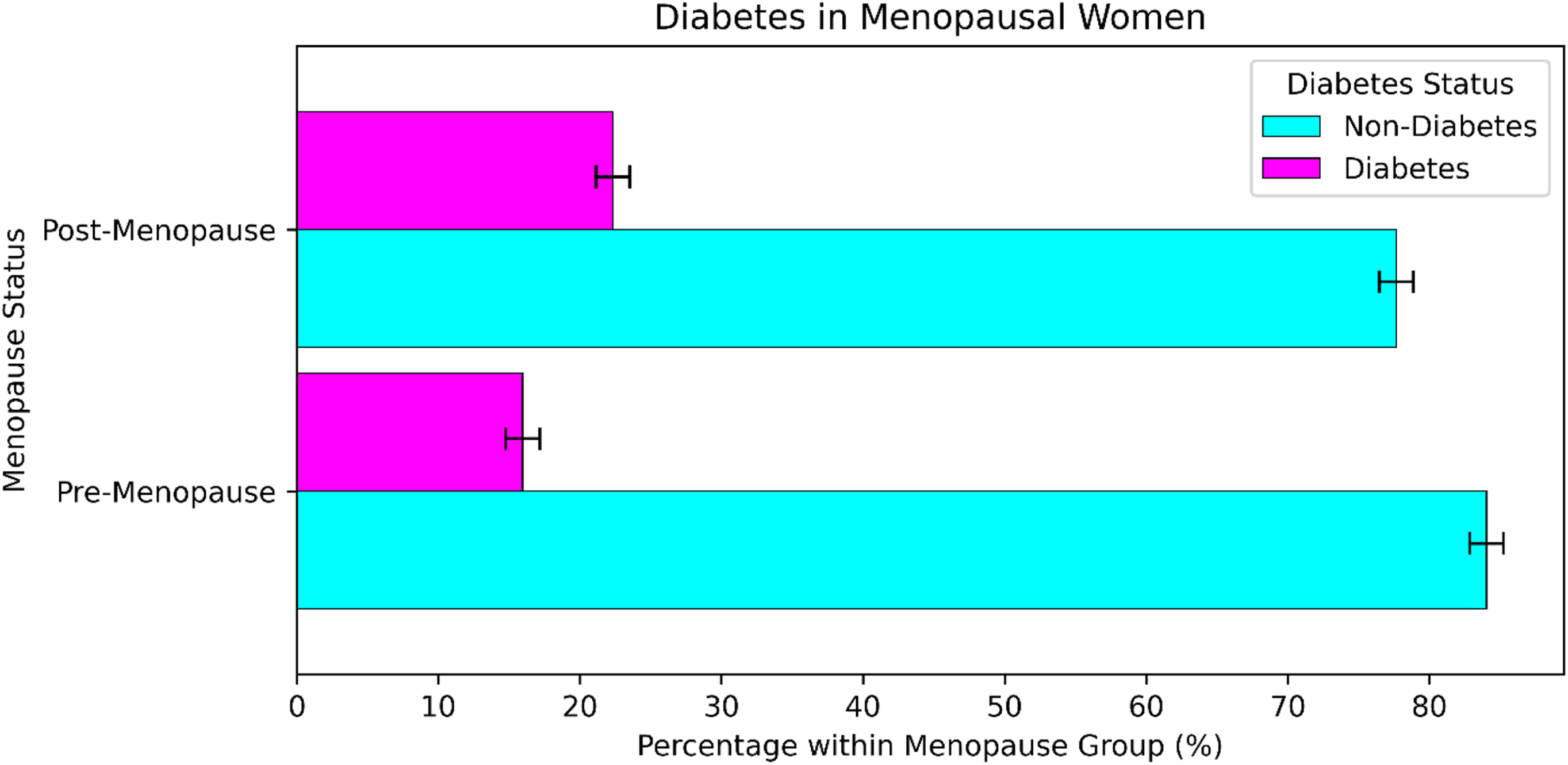
Diabetic and non-diabetic individuals evaluated amongst menopausal status.

In the post-menopausal category, 77.66% of female participants are non-diabetic and 22.34% of female participants are diabetic. The p value is equal to 0.0001 which indicates that the difference is statistically significant (p < 0.05). The standard error is 1.19.

### 3.9 Menopause and the Association with Nocturnal Dipping Patterns in Female Participants

The correlation between menopausal status and nocturnal dipping pattern in female participants is found in Figure 7(c). In the normal dipping category, 61.54% of female participants are post-menopausal women and 38.46% of female participants are pre-menopausal women with a p value of 0.56. The difference is not statistically significant (p > 0.05) and the standard error is 1.82.

**Figure 7(c).**
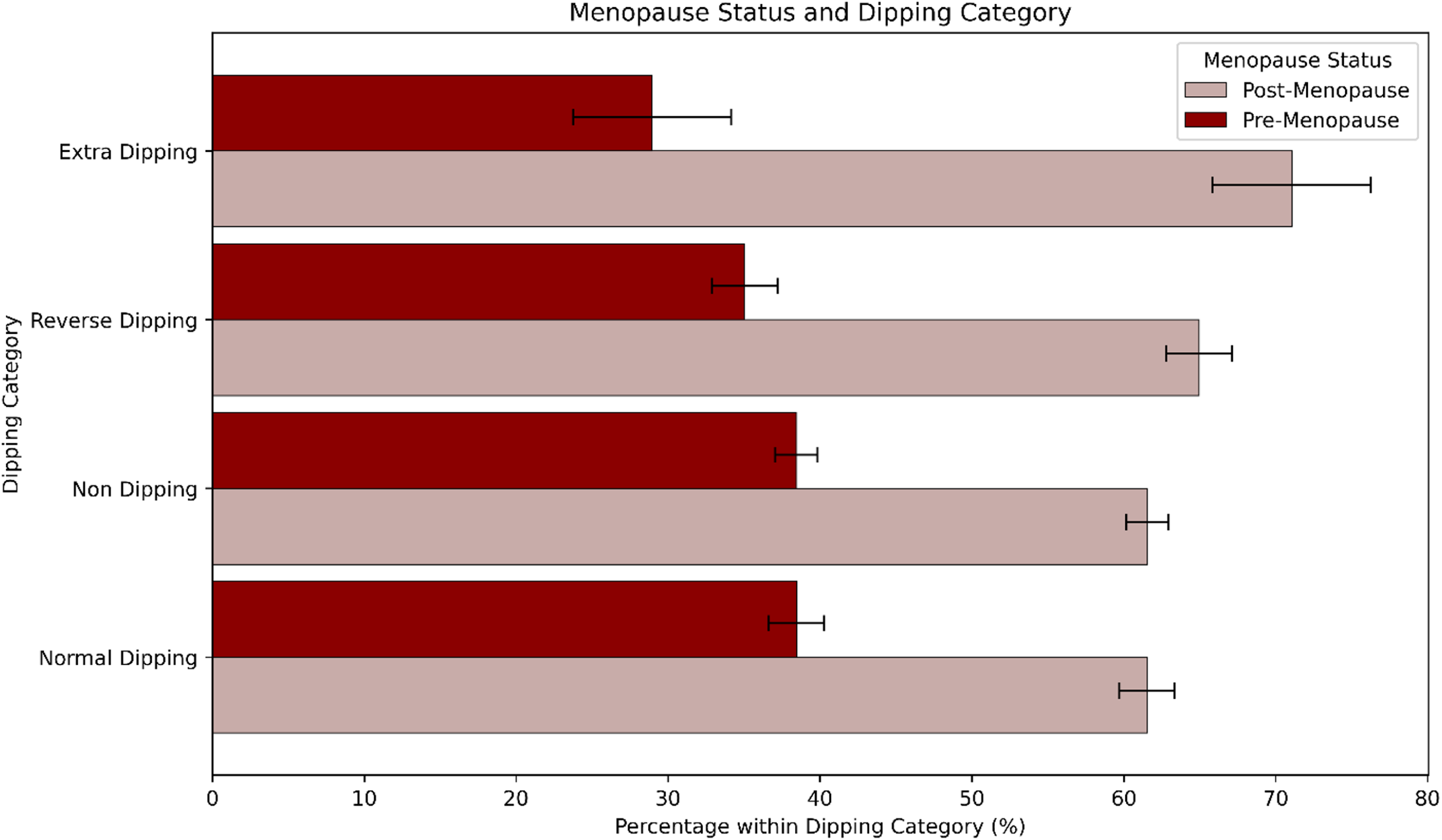
Post-menopausal and pre-menopausal women participants ated amongst nocturnal dipping patterns.

In the non-dipping category, 61.56% of female participants are post-menopausal women and 38.44% of female participants are pre-menopausal women with a p value of 0.36. The difference is not statistically significant (p > 0.05) and the standard error is 1.39.

For the female participants in the reverse dipping category, 64.96% are post-menopausal women and 35.04% are pre-menopausal women. The p value is equal to 0.23 which indicates that these results are not statistically significant (p > 0.05). The standard error is 2.16.

For the female participants in the extra dipping category, 71.05% are post-menopausal women and 28.95% are pre-menopausal women with a p value of 0.15. The difference is not statistically significant (p > 0.05) and the standard error is 5.20.

### 3.10 Menopause and the Association with Early Morning Hypertension in Female Participants

The correlation between menopausal status and early morning hypertension in female participants is found in Figure 7(d). Among the pre-menopausal category, 83.40% of female participants do not experience early morning hypertension and 16.60% of female participants experience early morning hypertension. The p value is equal to 0.0065 therefore the difference is statistically significant (p < 0.05). The standard error is 1.21.

**Figure 7(d).**
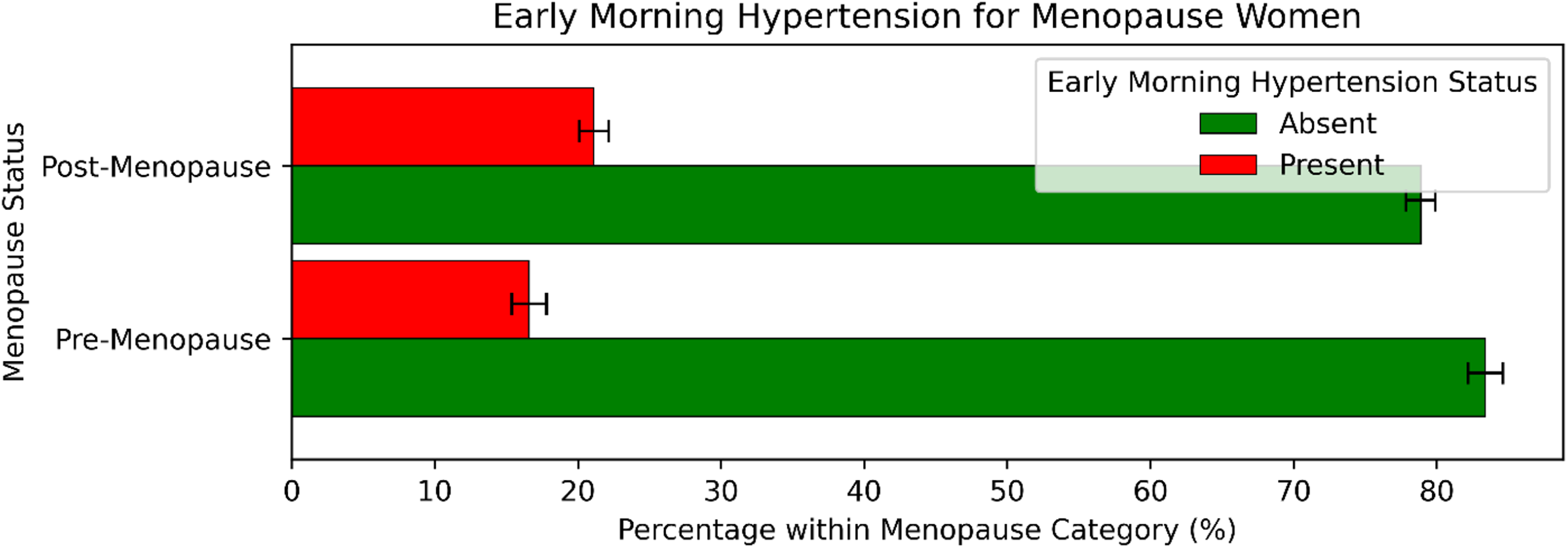
Menopausal status for women with and without early morning hypertension.

Among the post-menopausal category, 78.88% of female participants do not experience early morning hypertension and 21.12% of female participants experience early morning hypertension. The p value is equal to 0.0065 which indicates that these results are statistically significant (p < 0.05). The standard error is 1.03.

### 3.11 Early Morning Hypertension in Total Sample Population

Prevalence of early morning hypertension in the total sample population is found in Figure 8(a). There are 19.62% of participants that experience early morning hypertension and 80.38% of participants do not experience early morning hypertension. The SE is equal to 0.53.

**Figure 8(a).**
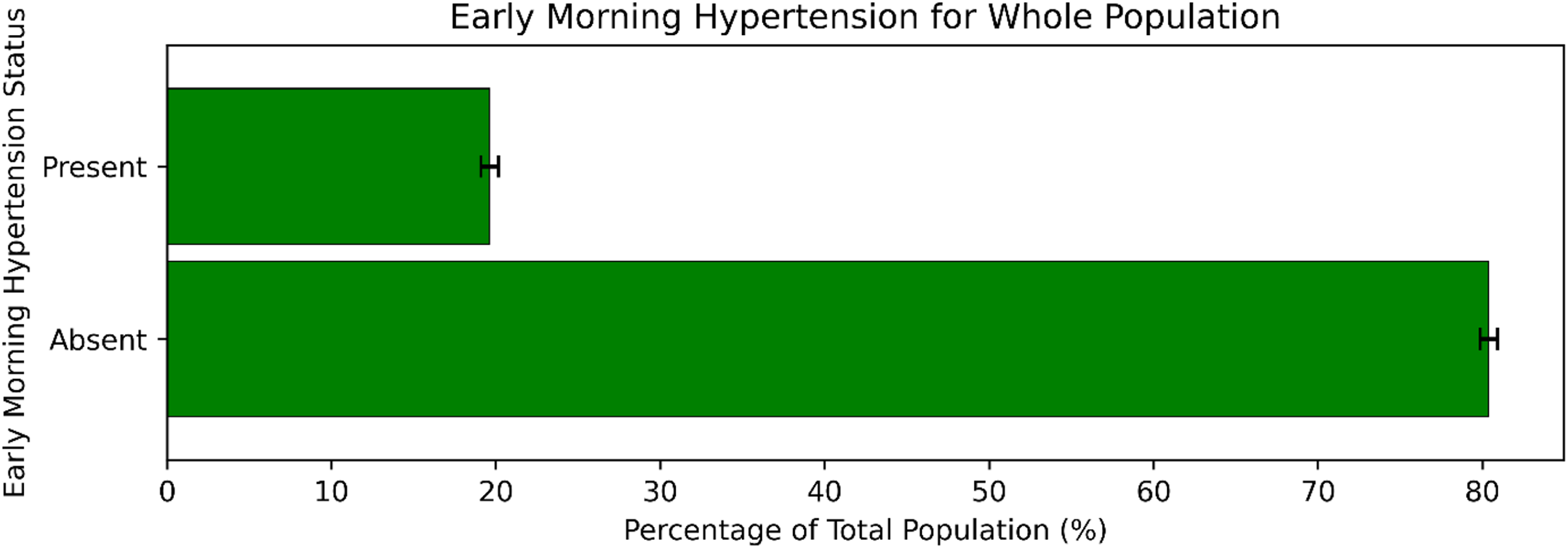
Participants with early morning hypertension and without early morning hypertension among the total sample.

### 3.12 Early Morning Hypertension in Association with Biological Sex

The correlation between early morning hypertension and biological sex is found in Figure 8(b). For male participants, 19.77% experience early morning hypertension and 80.23% do not experience it (SE = 0.71). For female participants, 19.43% experience early morning hypertension and 80.57% do not experience it (SE = 0.79). The p value is equal to 0.77, therefore the difference is not statistically significant (p > 0.05).

**Figure 8(b).**
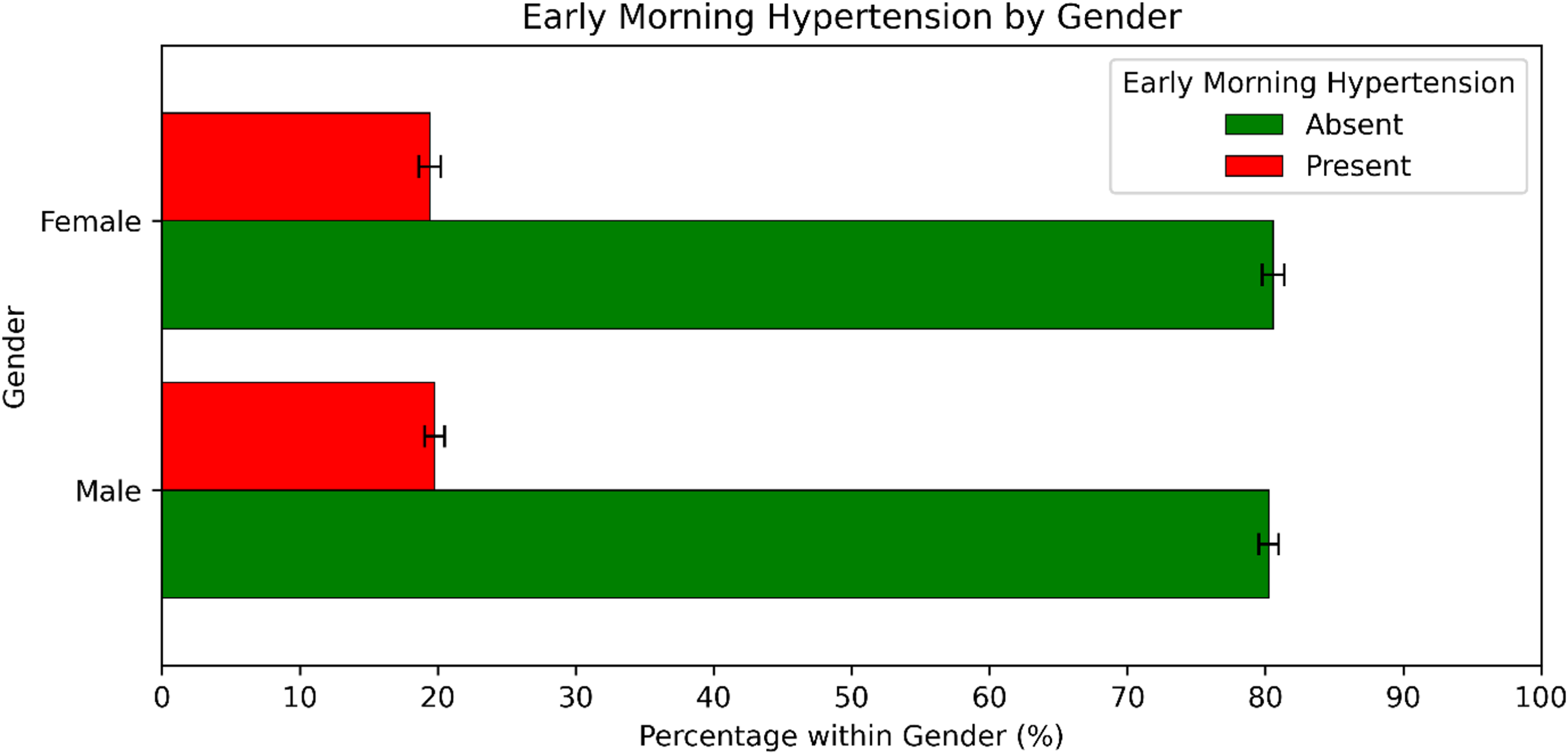
Male and female participants with and without early morning hypertension.

### 3.13 Intercomparisons of Blood Pressure and Heart Rate Data

In this study, systolic and diastolic blood pressure (BP) measurements, along with heart rate (HR), were recorded continuously throughout the day for all participants. The mean systolic BP was 125.16 ± 13.85 mmHg, while the mean diastolic BP was 74.63 ± 9.94 mmHg.

Systolic BP was further analyzed separately for daytime and nighttime periods. The mean daytime systolic BP was 129.37 ± 12.87 mmHg, while the mean nighttime systolic BP was 120.95 ± 14.82 mmHg.

Similarly, diastolic BP measurements were analyzed for daytime and nighttime periods. The mean daytime diastolic BP was 79.67 ± 9.42 mmHg, while the mean nighttime diastolic BP was 69.58 ± 10.46 mmHg.

Heart rate (HR) was recorded for both daytime and nighttime periods. The mean daytime HR was 78.71 ± 10.39 beats per minute, while the mean nighttime HR was 69.19 ± 10.36 beats per minute. An intra-comparison of daytime and nighttime HR values using a paired t-test revealed a statistically significant difference (p < 0.05).

## 4. DISCUSSIONS

This study highlights the vital role of ABPM in understanding nocturnal BP patterns and their associations with BMI, diabetes, and menopausal status. The findings reinforce ABPM’s value as a comprehensive tool for assessing cardiovascular risk and guiding effective management strategies.

### 4.1 Hypertension and Nocturnal BP Patterns

Our findings show that the extra dipping category is the only group in which the number of participants with hypertension exceeds those without, although this difference is not statistically significant (p > 0.05). Among non-hypertensive participants, the majority exhibit reverse dipping (63.06%) or non-dipping (60.77%) nocturnal blood pressure patterns. These abnormal dipping profiles have been associated with chronic conditions such as kidney disease, which is often marked by increased urinary protein excretion which is a strong predictor of future cardiovascular events, renal complications, and mortality [65]. Consequently, participants demonstrating reverse dipping and non-dipping profiles are therefore at heightened risk for progression to lifelong cardiometabolic conditions. Ambulatory blood pressure monitoring (ABPM) plays a critical role in identifying these patterns early and enabling the implementation of precision health interventions before irreversible damage occurs.

### 4.2 Hypertension and BMI

The overall trend observed in our findings indicates that hypertension prevalence increases with rising BMI, although the severely obese category is the only one with a statistically significant difference (p = 5.55e-15, p < 0.05). In the severely obese group, the number of hypertensive participants surpasses that of non-hypertensive participants, which is not seen in any other BMI category. This pattern suggests a correlation between excess adiposity and elevated blood pressure, a relationship well documented in existing literature. Proposed mechanisms underlying this association include sympathetic nervous system overactivation, stimulation of the renin-angiotensin-aldosterone system, dysregulation of adipose-derived cytokines, insulin resistance, and both structural and functional renal alterations in individuals with obesity [66].

### 4.3 Nocturnal BP Patterns and BMI

Non-dipping is the most prevalent nocturnal blood pressure pattern across all BMI categories, except in the underweight group, where reverse dipping is the most common. Our findings show non-dipping and reverse dipping patterns generally increase with higher BMI, while normal dipping and extreme dipping patterns decline. Previous research has established a direct relationship between the extent of nocturnal blood pressure reduction and a decrease in sympathetic nervous system activity [65]. Weight gain has been shown to activate the sympathetic nervous system with the degree of activation proportional to the percentage increase in body weight [67]. This heightened sympathetic activity may contribute to the blunting or reversal of normal nocturnal dipping patterns, as reflected in the increased prevalence of non-dipping and reverse dipping from our findings. The concurrent decline in extreme dipping patterns with rising BMI further supports this association. These observations are consistent with findings by Cuspidi et al., who reported that higher BMI is linked to a reduced nocturnal decline in blood pressure and a greater risk of both cardiac and extracardiac organ damage [68].

Collectively, these findings underscore the clinical importance of ambulatory blood pressure monitoring (ABPM), particularly for individuals with overweight or obesity, as early identification of abnormal dipping patterns may facilitate timely interventions through lifestyle changes and pharmacological therapies to prevent the development of associated comorbidities.

### 4.4 Nocturnal BP Patterns and Diabetes

The findings of our study concluded that among hypertensive individuals, the prevalence of diabetes was significantly higher compared to non-hypertensive participants (17.59% vs. 21.18%, *p* = 0.001), reinforcing the known comorbidity between elevated BP and impaired glucose metabolism seen in current hypertension literature [58]. While a majority of participants across all the dipping patterns were non-diabetic, a statistically significant association was found only within the normal dipping group (*p* = 0.02), where 17.78% of participants were diagnosed as diabetic. Though not markedly statistically significant, the non-dipping and reverse-dipping groups had modestly higher proportions of diabetic individuals, suggesting a trend towards disrupted circadian BP regulation amongst participants with diabetes. Relating to current hypertension literature, these patterns may reflect underlying autonomic dysfunction and metabolic disturbances that are prevalent in individuals with both diabetes and hypertension, further underscoring the importance of ABPM technology in uncovering these high-risk patient profiles [58].

### 4.5 Menopausal Status and Hypertension

In our study, 38.97% of the pre-menopausal women population are diagnosed with hypertension, compared to the 61.03% of post-menopausal women who are diagnosed with hypertension. Our findings show the correlation between menopausal status and hypertension. The difference was not statistically significant (p > 0.05). Within our non-hypertensive category, 63.46% of post-menopausal women and 36.54% of pre-menopausal women were non-hypertensive, though the difference is not statistically significant (p > 0.05). Our results indicate that hypertension is modestly more prevalent among pre-menopausal women in this sample. These findings challenge the traditional view that elevated BP primarily develops post-menopause, suggesting that younger women may already be exhibiting early risk factors relating to cardiovascular disease [69]. Current scientific literature has recently begun studying hypertension status in women across age groups, finding that the prevalence of hypertension has increased significantly in women of all age groups, due to possible rising rates of elevated BMI and obesity amongst women raising the risk for hypertension [70]. Possible hypertension medication usage in the post-menopausal women population and possible oral contraceptive pill usage in pre-menopausal women with contraindications for hypertension-related risks could possibly explain the data trend noted. Pre-menopausal women with hypertension are vulnerable to developing comorbid conditions such as diabetes, metabolic syndrome, and cardiovascular disease [71]. Both groups of pre-menopausal and post-menopausal women are at risk of developing hypertension. Therefore, ABPM use is critical to monitor BP throughout the day. Early detection in pre-menopausal women through ABPM can facilitate targeted interventions and treatment plans to help prevent the progression of disease and improve long-term outcomes.

### 4.6 Menopausal Status and Diabetes

As shown in our study findings, a higher proportion of postmenopausal women are diagnosed with diabetes (22.34%) compared to pre-menopausal women (15.96%). The decline in estrogen levels during menopause has been associated with increased insulin resistance which is a state in which the body’s cells respond less effectively to insulin, leading to elevated blood glucose levels and a greater risk of developing type 2 diabetes [72]. While the link between low estrogen and insulin resistance is well established, evidence regarding the relationship between menopause and diabetes remains mixed. Some studies suggest that age may act as a confounding factor, making it difficult to determine whether the increased diabetes risk is directly attributable to menopause itself or to age-related metabolic changes [73]. Nonetheless, the observed increase in diabetes among postmenopausal women underscores the importance of monitoring glucose regulation during and after the menopausal transition, particularly for individuals with additional metabolic risk factors associated with abnormal dipping patterns.

### 4.7 Menopausal Status, Nocturnal Dipping Patterns and Early-Morning Hypertension

As shown in our study findings, early morning hypertension is more prevalent among postmenopausal women (21.12%) compared to premenopausal women (16.60%), a difference that is statistically significant (p = 0.0065, p < 0.05).

Under normal conditions, blood pressure dips during sleep due to increased parasympathetic and decreased sympathetic nervous system activity [74]. However, research by Farag et al. found that postmenopausal women have reduced parasympathetic activity at rest, and therefore an increased sympathetic tone [75]. Our findings further shows that extreme dipping (71.05%) is the most common nocturnal BP pattern in postmenopausal women, followed by reverse dipping (64.96%). Elevated sympathetic activity has been associated with reverse dipping patterns which aligns our findings with the findings by Su et al., who linked menopause to increased rates of reverse dipping [76–77]. However, the predominance of extreme dipping in our sample contrasts with existing literature, which typically reports more blunted dipping patterns in postmenopausal women due to reduced parasympathetic tone [78]. Despite this discrepancy, our findings reinforce the clinical relevance of ambulatory blood pressure monitoring (ABPM) for postmenopausal women. Research suggests that those with exaggerated dipping and early morning hypertension may be at increased risk for both silent and clinical cerebral ischemia due to sharp morning BP surges. Early detection through ABPM may enable timely intervention and risk reduction in this population [79].

### 4.8 Nocturnal BP Patterns and Early-Morning Hypertension

In relation to early morning hypertension, our study found that 19.62% of the total study population experienced elevated BP within the first two hours of waking. Although early morning hypertension was observed at similar rates between males (19.77%) and females (19.43%), the differences were not statistically significant, thus indicating that early morning surges in BP are not sex-dependent in this cohort. The early morning hypertension surge is a clinically important period, as it is associated with an increased risk of acute cardiovascular events such as stroke and myocardial infarction. Prior clinical research emphasizes the association between morning BP surges and heightened incidence of stroke, myocardial infarction, and target organ damage [59]. These findings highlight the significance of ABPM testing in detecting early morning hypertensive episodes that may have gone otherwise unnoticed in standard office-based assessments.

### 4.9 ABPM and Clinical Implications

This study reinforces the importance of ABPM in detecting nocturnal hypertension and abnormal BP patterns that may be missed by traditional office-based BP measurements. By providing continuous BP readings over a 24-hour period, ABPM captures fluctuations during sleep, daily activities, and stress, offering a more comprehensive assessment of cardiovascular risk. Additionally, ABPM helps identify white coat and masked hypertension, reducing the risk of misclassification and improving diagnostic accuracy.

### 4.10 Target Outcomes and Recommendations

The findings emphasize the need to integrate ABPM into routine clinical practice to optimize hypertension management and improve patient outcomes. The ability of ABPM to detect nocturnal hypertension, evaluate BP variability, and monitor treatment effectiveness makes it an essential tool in cardiovascular care. This study advocates for the broader use of ABPM in both clinical and pharmacy settings, particularly for high-risk populations such as overweight individuals, diabetics, and postmenopausal women.

## 5. CONCLUSION

This study highlights the critical role of ABPM in assessing nocturnal BP patterns and their links to Body Mass Index (BMI), diabetes, and menopausal status to ultimately assess risk of adverse outcomes such as the development of cardiovascular complications. The findings revealed a significant prevalence of unhealthy non-dipping and reverse dipping BP patterns among overweight and obese individuals, stressing the importance of early detection and targeted interventions. Additionally, the study shed light on the complex connections between hypertension, diabetes, and menopausal status, with postmenopausal women showing a higher risk of experiencing early morning hypertension.

ABPM offers a more comprehensive approach to assessing cardiovascular risks compared to traditional office-based BP measurements. By capturing 24-hour BP fluctuations, ABPM provides greater diagnostic accuracy, including the ability to detect nocturnal hypertension, white coat hypertension, and masked hypertension. These capabilities support more precise treatment planning and improved hypertension management.

The results underscore the need for wider integration of ABPM into clinical and pharmacy settings to enhance patient care and cardiovascular health outcomes. Future research should focus on long-term trends in BP variability and evaluate how personalized interventions can reduce cardiovascular risks in high-risk populations.

Incorporating ABPM into routine practice allows healthcare providers to improve risk assessment, tailor treatment strategies, and ultimately reduce the global burden of hypertension and its associated complications.

## Data Availability

The data that support the findings of this study are not publicly available. However, the data and analysis will be made available to the editors and peer reviewers upon reasonable request during the review process.

